# Relationship Extraction for Adverse Drug Events in Clinical Notes Using Large Language Models

**DOI:** 10.64898/2026.05.28.26354362

**Authors:** Joseph M Plasek, Yiming Li, Mary G Amato, Dinah Foer, Diane L. Seger, Shayma Alzaidi, Huiyuan Zhou, Gretchen Purcell Jackson, David W Bates, Li Zhou

## Abstract

**Background:** Adverse drug events (ADEs) are a critical indicator of patient safety but are often documented only in free-text clinical notes. The potential of recent advances in natural language processing (NLP), particularly generative large language models (LLMs), to identify ADEs remains understudied. This study aimed to compare the performance of multiple LLMs in identifying ADE-Drug relationships in inpatient and ambulatory clinical notes.

**Methods:** We used clinical notes from the 2018 National NLP Clinical Challenge (n2c2) ADE dataset (inpatient; n=505) and from outpatient encounters (n=2,555) between October 1, 2018, and December 31, 2019, at a large academic medical center based in New England. Notes were pre-processed into snippets for model input. Evaluated Models included: GPT-4o, GPT-4o-mini, LLAMA 3.3-70B and their instruction fine-tuned variants (including low-rank adapters for LLAMA). Performance was assessed using both strict and relaxed evaluations (precision, recall, and F1) for all models, followed by manual evaluation (exact semantic match, partial match, missing ADE, drug mention only, not a drug, or wrong) of the two best-performing models.

**Results:** GPT-4o and GPT-4o-mini were the top-performing models among those evaluated. GPT-4o consistently outperformed GPT-4o-mini in ADE extraction across both datasets, with higher F1-scores (0.524 vs. 0.381) and a more balanced precision-recall profile. Both models captured ADEs effectively in explicit and complex clinical contexts, although limitations included misclassification of pre-existing allergies and occasional conflation of therapeutic indications with adverse effects. GPT-4o achieved higher exact match coverage and fewer errors across clinical notes, indicating more reliable performance in both inpatient and ambulatory settings.

**Conclusion:** This work establishes a foundation for integrating LLM methods into real-world drug safety surveillance, with direct implications for improving patient safety.

## Introduction

The annual economic impact of treating adverse drug events (ADEs) in the United States is estimated to be $1.56-$5.6 billion in the inpatient setting (1) plus $8 billion in the ambulatory setting (2). A multicenter retrospective review of available electronic health records (EHR) data for a random sample of 2,809 inpatient admissions found 13.6 ADEs per 100 admissions (3); and among 3103 ambulatory patients, 1.25 ADEs per 100 encounters (4,5). Additional details from the ambulatory arm of this study suggested that over a one-year period, 5% of patients experienced an ADE (mean of 4 encounters per patient)(5). We previously estimated the overall prevalence of ambulatory clinical notes containing ADEs to be 1.97 per 100 patient encounters (or 2.52 per 100 encounters when considering possible ADEs) (6).

ADEs are defined as any injury resulting from medical interventions related to a drug (7). Rapid detection of ADEs in patient care settings is important so that patients can receive prompt treatment to reduce the duration of harm. Steps can be taken to reduce the frequency of preventable medication related to patient harm. However, many traditional methods used to identify ADEs are either inaccurate, labor intensive or inefficient. Most ADEs are not reported to spontaneous reporting systems such as the US Food and Drug Adverse Event reporting system or health system incident reporting systems and use of billing codes from claims data may lack sensitivity (7). Moreover, free-text clinical notes, which may contain important descriptions of ADEs, are not assessed by routine data monitoring systems that rely on structured data or reporting tools (8,9). This exclusion prevents pharmacovigilance efforts focused on early ADE detection (e.g., to find medication prescribing errors to prevent ADEs or identify ADEs early to reduce duration or severity of harm) or prediction (e.g., to predict the therapeutic response to medications, to balance therapeutic benefit with ADE risk, to guide the selection of safe and effective medications) (10).

Named entity recognition (NER), which involves labeling text segments as Drug or ADE, was the initial focus of natural language processing (NLP) approaches to ADE extraction. However, prior challenges have shown that top-performing systems achieve over 89% F1-measure on drug labeling, indicating that further improvements in NER offer limited benefit and would largely require overfitting to dataset-specific noise (11). Consequently, framing ADE extraction as a relationship classification task determining whether a given pair of ADE and Drug entities share a causal relationship has emerged as a more clinically informative approach (12,13). Chen et al. developed attention-based bidirectional long-short term memory (LSTM) and convolutional neural network (CNN) models for relation classification (14); Li et al. leveraged Bidirectional long-short term memory (BiLSTM) architectures with word-, part-of-speech–, and position-based embeddings (15); and Yang et al. applied classical machine learning approaches such as support vector machines (SVMs) and random forests using handcrafted entity, context, distance, and semantic features (16). As summarized in a recent comprehensive review by Li et al., these approaches span a wide range of model families, including SVMs, random forests, CNNs, BiLSTM variants, and Bidirectional Encoder Representations from Transformers (BERT)-based architectures. However, these methods were typically trained and evaluated under sentence-level or narrowly defined context settings, relied heavily on handcrafted features or task-specific supervision, and struggled to model cross-sentence ADE-Drug relations or handle cases involving multiple co-occurring drugs. These limitations constrain generalizability across heterogeneous EHR documentation styles and motivate the need for approaches that can flexibly reason over variable-length clinical context while reducing dependence on rigid feature engineering (12). Silverman et al. adapted a clinical BERT model (UCSF-BERT) to identify serious adverse events (SAEs) within the specific context of 928 outpatient inflammatory bowel disease (IBD) notes. UCSF-BERT achieved the highest accuracy (88–92%) and macro F1 (61–68%) in extracting drug–SAE pairs compared to eight other candidate models. While these results significantly outperformed prior approaches in identifying hospitalization events emergent to medication use, the authors noted the focus on this specific outpatient subset when evaluating broader generalizability (17). To overcome these challenges, recent advances in generative large language models (LLMs), which are trained on vast amounts of textual data and exhibit strong language comprehension capabilities, provide a promising framework for improving the identification of ADEs and enhancing patient safety (18–20).

Koon et al. demonstrated that state-of-the-art generative LLMs can achieve performance comparable to fine-tuned transformer models for ADE-Drug relation prediction in discharge summaries, particularly excelling as recall-optimized models when contextual information is preserved. Their work further suggests that hybrid pipelines combining fine-tuned models with generative LLMs can balance performance and computational efficiency (21).

Prior annotated NLP benchmark datasets have examined ADEs in FDA drug labels, inpatient clinical notes, oncology notes, and social media data sources, but have not systematically investigated ADEs in the ambulatory setting, where documentation is shorter, noisier, and more heterogeneous (22–28). Moreover, the interaction between context windowing strategies and LLM performance in real-world EHR text remains underexplored.

## Objectives

The objective of this study was to compare the performance of different LLMs in identifying ADE-Drug relationships in clinical notes from inpatient(23) and ambulatory(6) settings.

## Methods

### Study Design and Oversight

We employed a retrospective cross-sectional study design (Figure 1) (6,23). This study was approved by the Mass General Brigham Institutional Review Board (IRB reference number: 2019P002998). Informed consent from patients was exempted by the IRB, as only de-identified retrospective data were used and where results are only presented in aggregate. We followed the AI reporting guidelines from TRIPOD AI (29), MI-CLAIM (30), and NLP reporting quality guidelines from Davidson et al. (31) where applicable.

**Figure 1.**
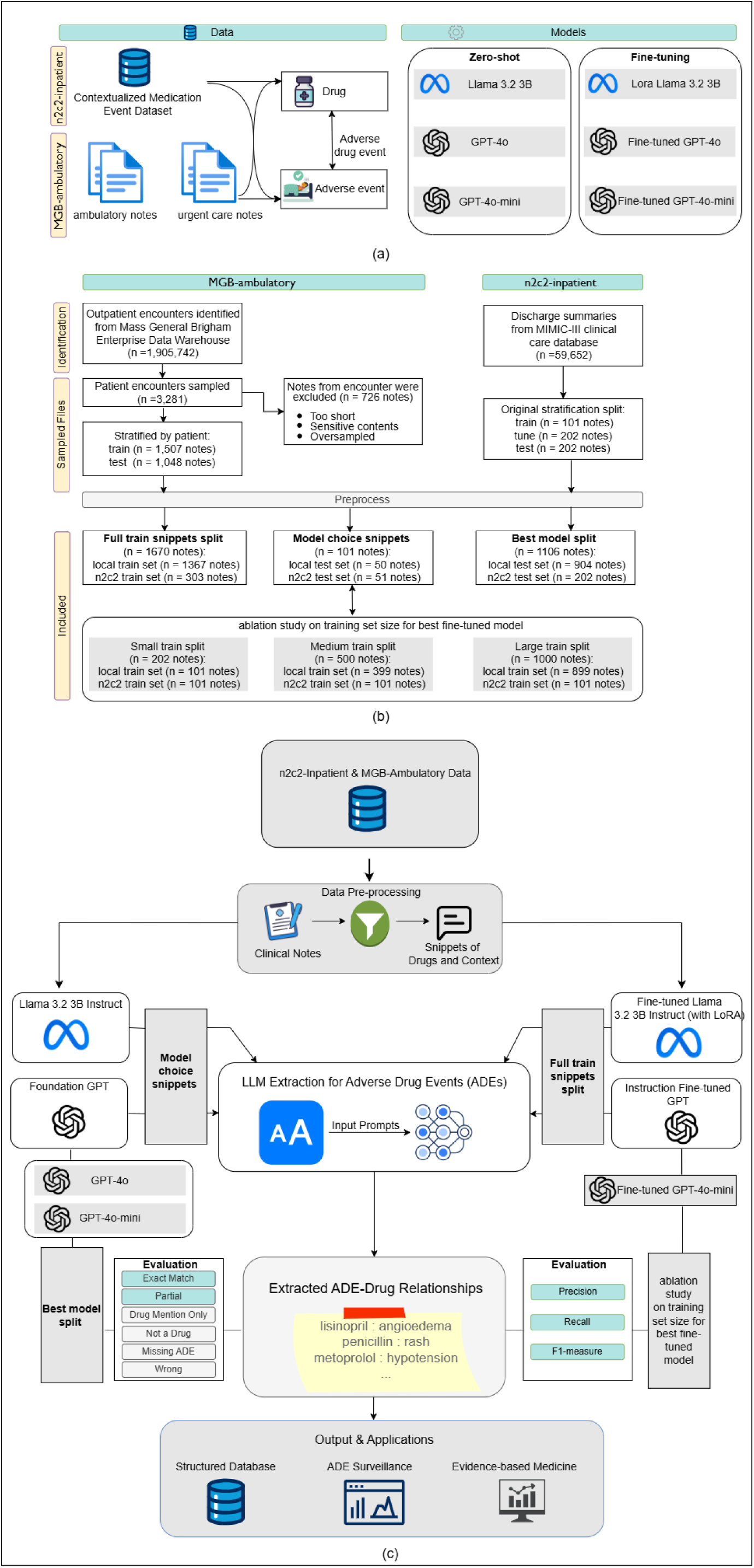
Overview of datasets, cohort selection, and workflow for LLM-based adverse drug event (ADE) extraction. (a) The study utilized two data sources: the n2c2-inpatient 2018 Contextualized Medication Event dataset and a local clinical corpus comprising ambulatory and urgent care notes from the Mass General Brigham health system. Two types of labels were used for evaluation: drug and adverse drug event (ADE). Three large language models (LLMs) were evaluated in both zero-shot and fine-tuned settings: LLaMA 3.2B, GPT-4o, and GPT-4o-Mini. **(b)** Cohort selection and data stratification for both corpora. From the local corpus, 3,281 outpatient encounters were sampled, stratified into training, tuning, and test sets by patient, with exclusions made for sensitive content. The n2c2-inpatient dataset was used as released, with predefined train/test splits. Snippets were split into subsets for model selection, best model evaluation, and ablation studies assessing the impact of training set size. **(c)** End-to-end workflow for LLM-based ADE extraction. Clinical notes were preprocessed into input snippets highlighting drugs and their surrounding context. LLMs (both zero-shot and fine-tuned) extracted ADE relations using prompt-based queries. Output ADEs were manually evaluated using multiple criteria (e.g., exact match, partial, wrong), and performance was quantified using precision, recall, and F1-score. Final outputs were structured for downstream applications, including ADE surveillance and evidence-based medicine.

### Setting and Datasets

The two datasets used in this study are described in prior publications by Plasek et al and Henry et al. (6,23) Plasek et al described the prevalence and characteristics of ADEs in a cross-sectional study of 2,882 unique patients who had an outpatient encounter between October 1, 2018 through December 31, 2019 at any Mass General Brigham facility (e.g., primary care clinic, urgent care, allergy clinic) affiliated at the department level with Brigham and Women’s Hospital, Brigham and Women’s Faulkner Hospital, or the Brigham and Women’s Physicians Organization (6). Plasek et al used a stratified sampling strategy based on ICD-10 codes grouped by likelihood to create a balanced dataset (6,32).

MIMIC-III (Medical Information Mart for Intensive Care-III) consists of 59,652 discharge summaries for critical care admissions between 2001 to 2012 at Beth Israel Deaconess Medical Center (33). Henry et al described the sampling and stratification of the 505 MIMIC-III discharge summaries identified by ICD code descriptions that were used in the training or evaluation of multiple NLP tools as part of the 2018 n2c2 shared task on ADEs and medication extraction (23). To ensure a robust evaluation of model specificity, these negative samples from both datasets were distributed across the training and testing sets using stratified random sampling.

For both datasets, the annotations were designed to capture ADE–Drug relationships. Each annotated note contains explicit Drug entities and ADE entities, and the relationship between them was confirmed by human annotators according to a predefined schema. Detailed information about the annotation schema and entity/relation types is described in Plasek et al. and the original n2c2 publication (34,35).

### Measures

Gold standard relationships from ADE to Drug were annotated at the entity-relationship level in both datasets using similar definitions for the defining both the drug entity, ADE entity, and their relationship(6,23). In contrast to Plasek et al, adverse drug withdrawal events (ADWEs, n = 95) were included and re-coded as ADEs for this study. We only considered active or possible modifiers for the relationships in all metric calculations to reflect the correct identification of patient safety events, with other attributes being counted as false positives.

### Preprocessing

Due to the annotation guidelines, most of the annotated relationships in the datasets would be considered in-context (search in the immediate area around the drug name) instead of long-range context (consider linking ADEs and drugs across a long document/span). Our initial experimentation suggested that current LLMs are not well adept to long noisy input, which only seeks to inflate the token usage (and cost) for the LLMs, and exhibited degraded performance. We built a rule-based preprocessing pipeline based on the annotated gold standard to remove unnecessary text (i.e., a 225-character window on either side of an annotated relationship span - or entity where no relationship exists) and concatenated the resulting snippets. This pre-processing step allows us to effectively remove the structured sections of the note that do not contain ADE-related information. Note: as the structured medication and allergy sections were not labeled by the annotators in the MGB-ambulatory dataset due to differences in the annotation guidelines, these were effectively dropped as part of the pre-processing step, though they are prevalent in the n2c2-inpatient dataset. The models were not provided with the gold standard annotated concepts for Drug or ADE, just the free-text itself (several sentences worth, at minimum).

### Models

We experimented with multiple prompts and prompting strategies but opted to run the final comparison for this study using a simple prompt that seemed most likely to produce the desired output format. The prompt used for all models was: “system: “You are a pharmacist trained in chart review who is an expert at extracting ADE-Drug relationships. Please list output in the format ’drug | adverse reactions’. If no ADE-Drug relationships, do not print anything.”

We selected three models: GPT-4o, GPT-4o-mini, and Llama 3.2 3B based on availability and feasibility to run in a low resource environment or natively within the EHR (i.e., the model’s vendor is supported by our EHR vendor). GPT is a commercial product from OpenAI and is used in a HIPAA-compliant Microsoft Cognitive Services environment (36). Llama is an open source model from Meta that we ran in our local GPU server environment (4x NVIDIA L40s GPUs).

We fine-tuned the Llama model using Low-Rank Adaptation (LoRA) with the default hyperparameters (i.e., r = 64, lora_alpha=32, lora_dropout = 0.1, bias = ‘none’, target modules = [‘q_proj’,’k_proj’,’v_proj’,’o_proj’], 50 virtual tokens, 250 max_seq_length). We instruction fine-tuned both GPT models with a temperature of 0, top p of 0.85, and max tokens 800.

### Statistical Analysis

We extended that sampling strategy in Plasek et al to randomly sample within each strata to create a train and test set (34). We report descriptive statistics for the MGB-ambulatory train and test datasets before pre-processing. For the n2c2-inpatient corpus, the original splits for train (n = 101 clinical notes), tune (n = 202 clinical notes), and test (n = 202 clinical notes) were used (23). We report note length before and after pre-processing at the source level. We report the prevalence of notes containing at least one ADE.

The output for the generative models is designed to return human-readable sentences, rather than structured output. While we can prompt and fine-tune for structured output in a particular format, our initial experiments demonstrated that there are limits to the complexity of the output that can be asked for. Given that the context windows for some of the candidate generative LLMs are smaller than a full-length clinical note, getting accurate token spans for citations is not possible with the generative AI models. It is possible (likely) that the generative LLMs may also only return one representative if the same relationship is duplicated in the text. Thus, document-level performance metrics that ignore the span would be most suitable for this task. Model performance was evaluated via manual review using a relaxed lenient keyword in-context schema at the document level where medications identified as participating in an adverse drug reaction by the LLM were classified by reviewers as one of six different levels of correctness: an exact semantic match, partial march, missing ADE, drug-only mention, not a drug, or wrong/incorrect (Table 1). Two clinical pharmacists independently performed the manual review and classification, and a senior pharmacist adjudicated all conflicts until consensus was reached with inter-rater agreement being assessed using Cohen’s Kappa. The reviewers were blinded to model identity, ensuring unbiased evaluation. Performance metrics are reported in terms of precision, recall, and F1-score (Table 3). An additional sub-analysis was conducted on the ambulatory dataset to discern performance metrics by level of acuity (i.e., urgent care vs primary/specialty care). To quantify uncertainty, 95% confidence intervals (CIs) for precision, recall, and F1-score were estimated using nonparametric bootstrap resampling.

**Table 1.**
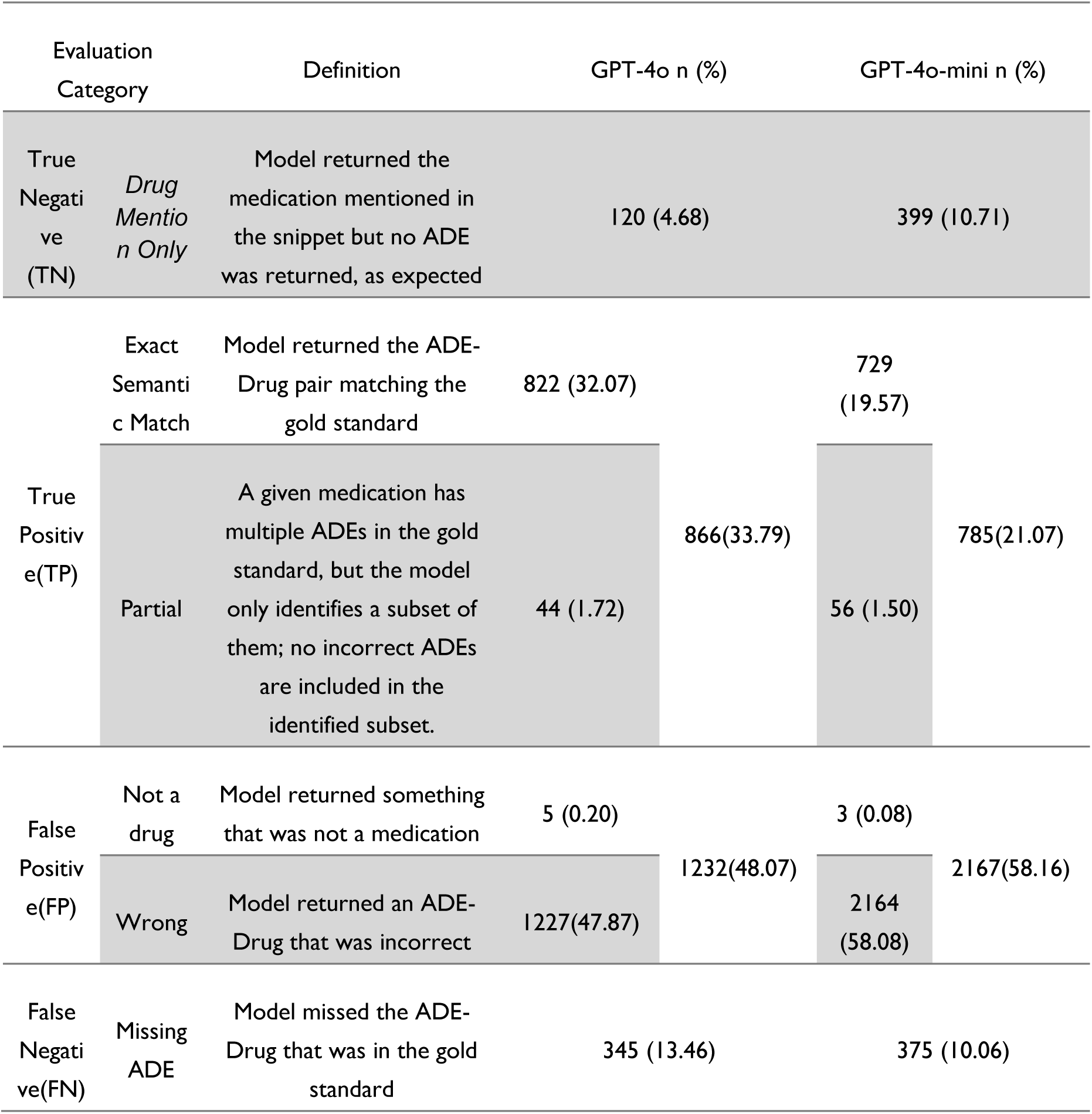
Evaluation schema and results for predicted Drug-ADE relations.

As model predictions were paired on the same documents, statistical differences in performance between models were assessed using McNemar’s test, which is appropriate for comparing paired nominal outcomes. All statistical tests were two-sided, and statistical significance was assessed at an α level of 0.05. We conducted an error analysis to identify trends.

## Results

In this section, we first describe the dataset characteristics, then we provide details on LLM selection, next we report our main results related to LLM performance of the top two LLMs, and finally provide an error analysis.

### Dataset Characteristics

Demographic characteristics for the train and test set of the MGB-ambulatory data set prior to pre-processing are available in the supplement (Supplement Table 1); the training set included 1,507 clinical notes and the test set included 1,048 clinical notes. The median number of tokens before and after pre-processing was 1683 and 390 in the MGB-ambulatory dataset and 3579 and 1769 in the n2c2-inpatient dataset, respectively (Table 2). The MGB-ambulatory corpus included 2,555 clinical notes before preprocessing (mean 1,924 tokens, range 15–11,735) and 2,322 clinical notes after preprocessing (mean 464 tokens, range 16–4,450). Of the 1048 notes in the test split, 450 contained at least one ADE, 417 had a drug mention but no ADE mention, and 181 were filtered out in the pre-processing step as they did not have a drug mention or ADE mention.

**Table 2:** Dataset Size Before and After Pre-Processing^&^.

|  | Number<br>of<br>Clinical<br>Notes | Mean<br>Tokens | Median<br>Tokens | Range<br>Tokens<br>[min,<br>max] |
| --- | --- | --- | --- | --- |
| Local before pre-processing* | 2555 | 1924.05 | 1683 | [15,<br>11735] |
| Local after pre-processing* | 2321 | 463.85 | 390 | [16,<br>4450] |
| n2c2 before pre-processing | 505 | 3813.53 | 3579 | [218,<br>13403] |
| n2c2 after pre-processing | 505 | 1911.72 | 1769 | [91,<br>5728] |

\* includes train and test splits only
&Notes after pre-processing contain at least one drug in the text.

**Table 3.**
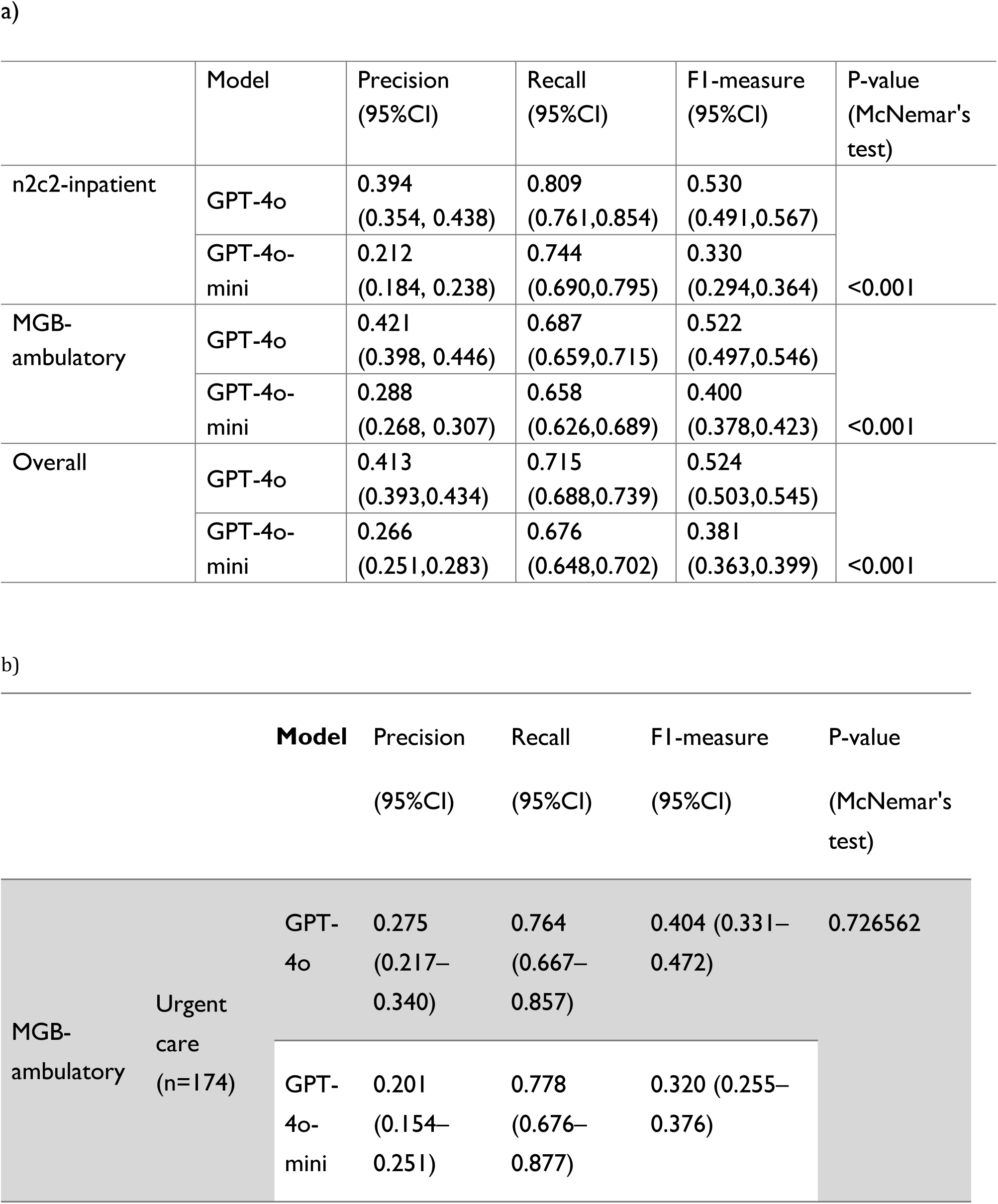
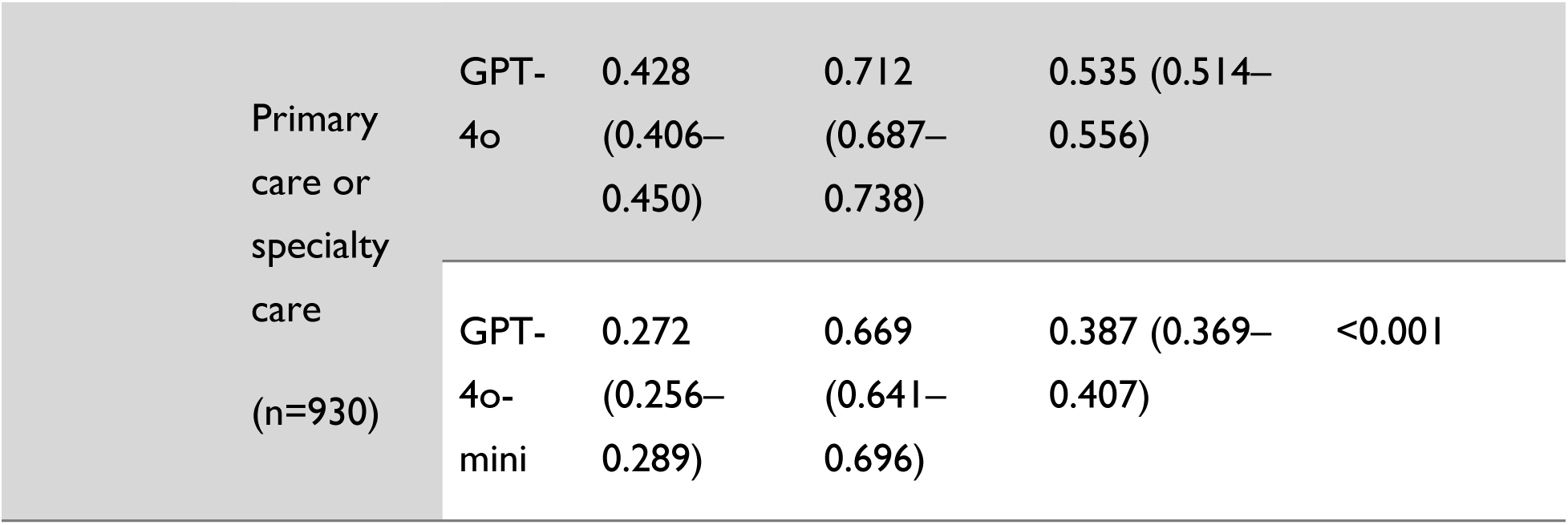
Performance comparison of GPT-4o and GPT-4o-mini a) across clinical datasets.

The n2c2-inpatient dataset contained 505 clinical notes, with mean tokens decreasing from 3,814 (range 218–13,403) before preprocessing to 1,912 (range 91–5,728) after preprocessing.

Of the 202 notes in the test set, 155 contained at least one ADE and the remaining 47 notes had at least one drug annotated but no ADEs mentioned.

### Model Selection

The 101 notes used as the model choice test set were randomly selected from the n2c2 (51 notes) and local (50 notes) datasets, of which 33 had at least one ADE and the remaining 68 notes had at least one drug annotated but no ADE mentioned. Two clinical research pharmacists with extensive experience detecting ADEs independently reviewed the output of the models using a randomized smaller sample of the dataset. Inter-rater agreement between the two pharmacists was calculated, with Cohen’s κ = 0.851, reflecting substantial agreement. After reviewing each sample dataset snippet against the gold standard and outputs from each model, they selected two candidates whose output was consistent with what they would expect of this task and that made the least mistakes. The first model selected was a small foundation model, GPT-4o-mini, selected because it had the highest combined “exact match” and “partial match” responses. The second model selected was a larger foundation model, GPT-4o, as it had the lowest combined total of missed ADE, not a drug, drug only, and wrong. The LLAMA models were non-performant, as they did not consistently produce the desired output format (instead, they largely started generating new clinical notes or reporting a summary of potential ADEs for a drug based on its knowledgebase, rather than returning the specific ADEs mentioned in the document), resulting in low relaxed-match F1 scores (0.0509–0.0552; Supplement Table 2). In the ablation study on fine-tuned training set size, we found that the fine-tuned model that performed the best was GPT-4o-mini with 1000 training examples which had the highest computed recall; however, it was not chosen due to degraded relaxed precision of 0.1004 (95%CI: 0.0639- 0.1507) (Supplement Table 2).

### Evaluation on n2c2-inpatient and MGB-ambulatory Ambulatory Notes’ Test Dataset

Table 1 presents the evaluation schema and comparative results for ADE-Drug relation extraction between GPT-4o and GPT-4o-mini. Six outcome categories were defined, encompassing correct predictions (exact semantic match, partial match), errors of omission (missing ADE), and errors of commission (drug-only mention, non-drug prediction, wrong relation). GPT-4o achieved a substantially higher proportion of exact semantic matches compared to GPT-4o-mini (32.1% vs. 19.6%), reflecting superior accuracy in identifying gold-standard ADE-Drug pairs. GPT-4o-mini generated more incorrect predictions overall, with nearly three-fifths of its outputs categorized as wrong (58.1% vs. 47.9% for GPT-4o). Both models showed relatively low proportions of partial matches (≤1.7%) and non-drug predictions (<0.3%). Although missing ADEs were observed at similar frequencies across models (13.5% for GPT-4o vs. 10.1% for GPT-4o-mini), GPT-4o demonstrated fewer spurious drug-only outputs (4.7% vs. 10.7%).

Overall, GPT-4o provided more reliable ADE-Drug relation extraction with fewer major errors and stronger alignment to the gold standard.

Table 3 presents performance characteristics of GPT-4o and GPT-4o-mini on adverse event extraction from the n2c2-inpatient and MGB-ambulatory datasets, as well as their combined evaluation. Across datasets, GPT-4o consistently outperformed GPT-4o-mini, with overall F1 of 0.524 (95%CI: 0.503-0.545) compared to 0.381 (95%CI:0.363-0.399). Both models achieved a higher recall than precision (e.g., GPT-4o overall recall 0.715 vs. precision 0.413), indicating a stronger tendency to capture adverse events at the expense of precision. These results indicate that GPT-4o provides more balanced and reliable performance across both inpatient and ambulatory settings compared with GPT-4o-mini. Within the MGB ambulatory cohort, further stratification by clinical setting revealed heterogeneity in model performance. In primary care or specialty care encounters (n = 930), GPT-4o demonstrated markedly superior performance to GPT-4o-mini, particularly in precision and F1-measure (0.535 vs. 0.387), with the difference remaining statistically significant (p < 0.001). In contrast, in the Urgent Care setting (n = 174), although GPT-4o achieved a higher F1-measure than GPT-4o-mini (0.404 vs. 0.320), GPT-4o-mini outperformed GPT-4o in recall.

Figure 2 presents a comparative evaluation of GPT-4o and GPT-4o-mini on an adverse drug event (ADE) relation extraction task, using two evaluation tags: “Exact Match” (both drug and ADE are correctly identified and linked) and “Partial” (Model returned the Drug, but the ADE returned was only partially correct). In the n2c2-inpatient corpus (Figure 2a), GPT-4o achieved 222 exact matches compared to 179 for GPT-4o-mini.

**Figure 2.**
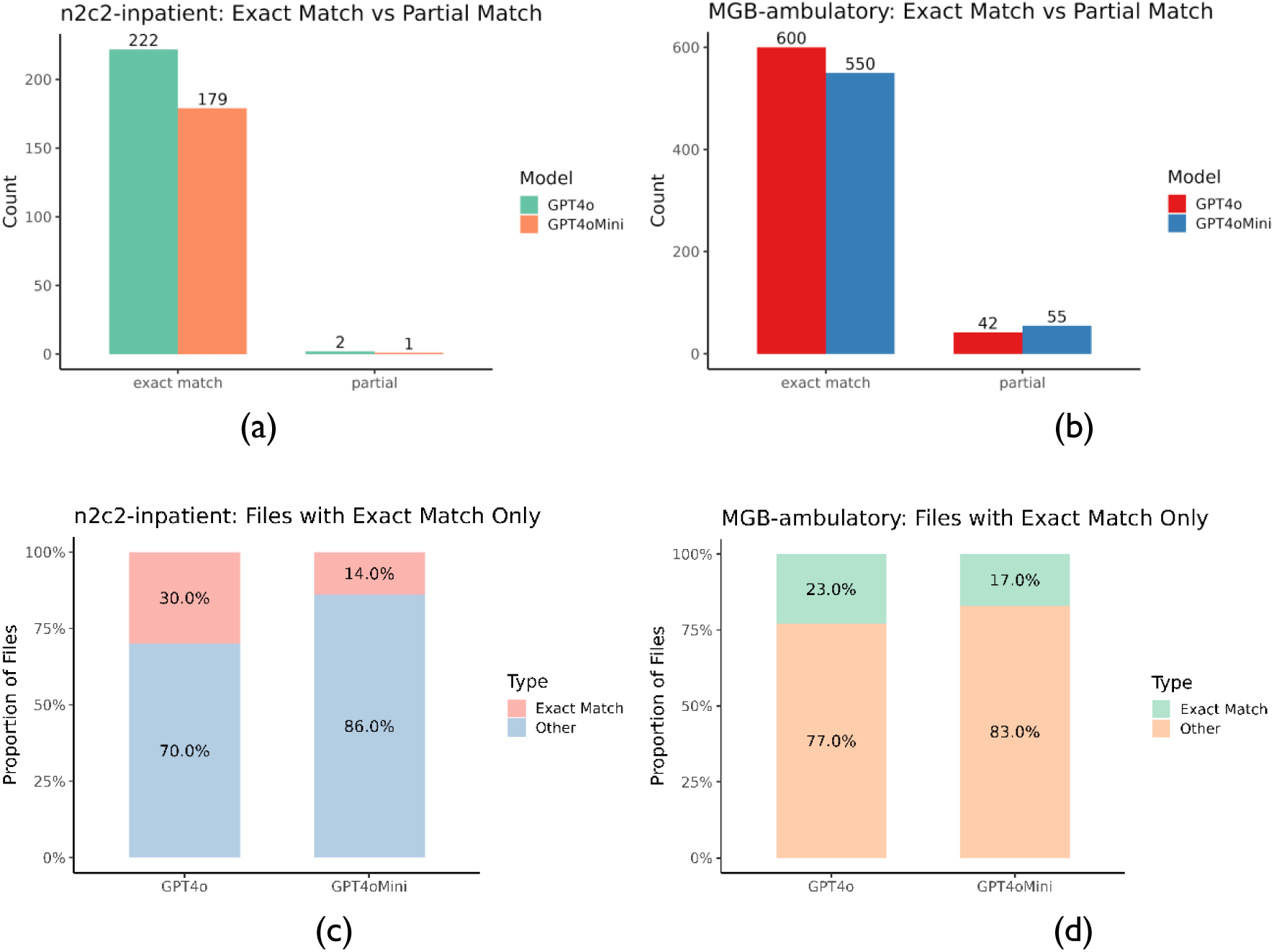
Performance comparison of GPT-4o and GPT-4o-Mini in adverse drug event (ADE) relation extraction. (a) shows the count of “Exact Match” and “Partial match” evaluation tags for GPT-4o and GPT-4o-Mini on the n2c2-inpatient corpus. (b) shows the same evaluation counts on the MGB-ambulatory corpus. (c) displays the proportion of clinical notes in the n2c2-inpatient corpus that contain only “Exact Match” evaluations, indicating high-fidelity predictions, while (d) shows this proportion for the MGB-ambulatory corpus.

The “Partial” tag was rarely used for both GPT-4o and GPT-4o-mini outputs. In the MGB-ambulatory corpus (Figure 2b), GPT-4o yielded 600 exact matches versus 550 for GPT-4o-Mini. Notably, GPT-4o again demonstrated fewer “Partial” instances (42 vs 55), indicating better end-to-end understanding of clinical relationships in unseen, real-world data. Figures 3c and 3d show the clinical note-level distribution of exact match performance. In the n2c2-inpatient corpus (Figure 2c), GPT-4o produced exact match–only evaluations in 30.0% of clinical notes, while GPT-4o-mini achieved this in only 14.0%. In the MGB-ambulatory corpus (Figure 2d), GPT-4o exhibited inferior performance, with 23.0% of clinical notes containing only exact matches, compared to 17.0% for GPT-4o-mini.

**Figure 3.**
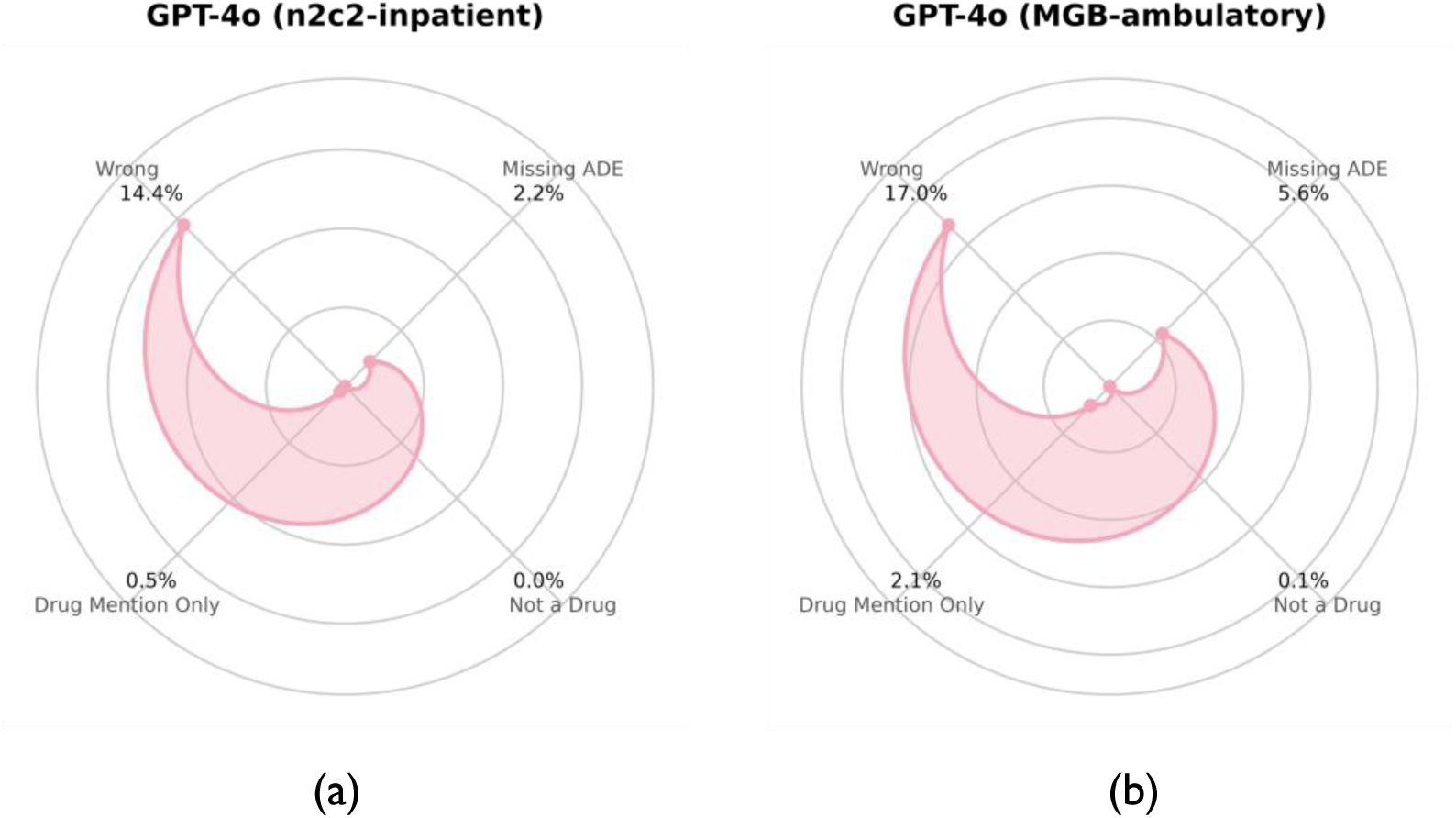
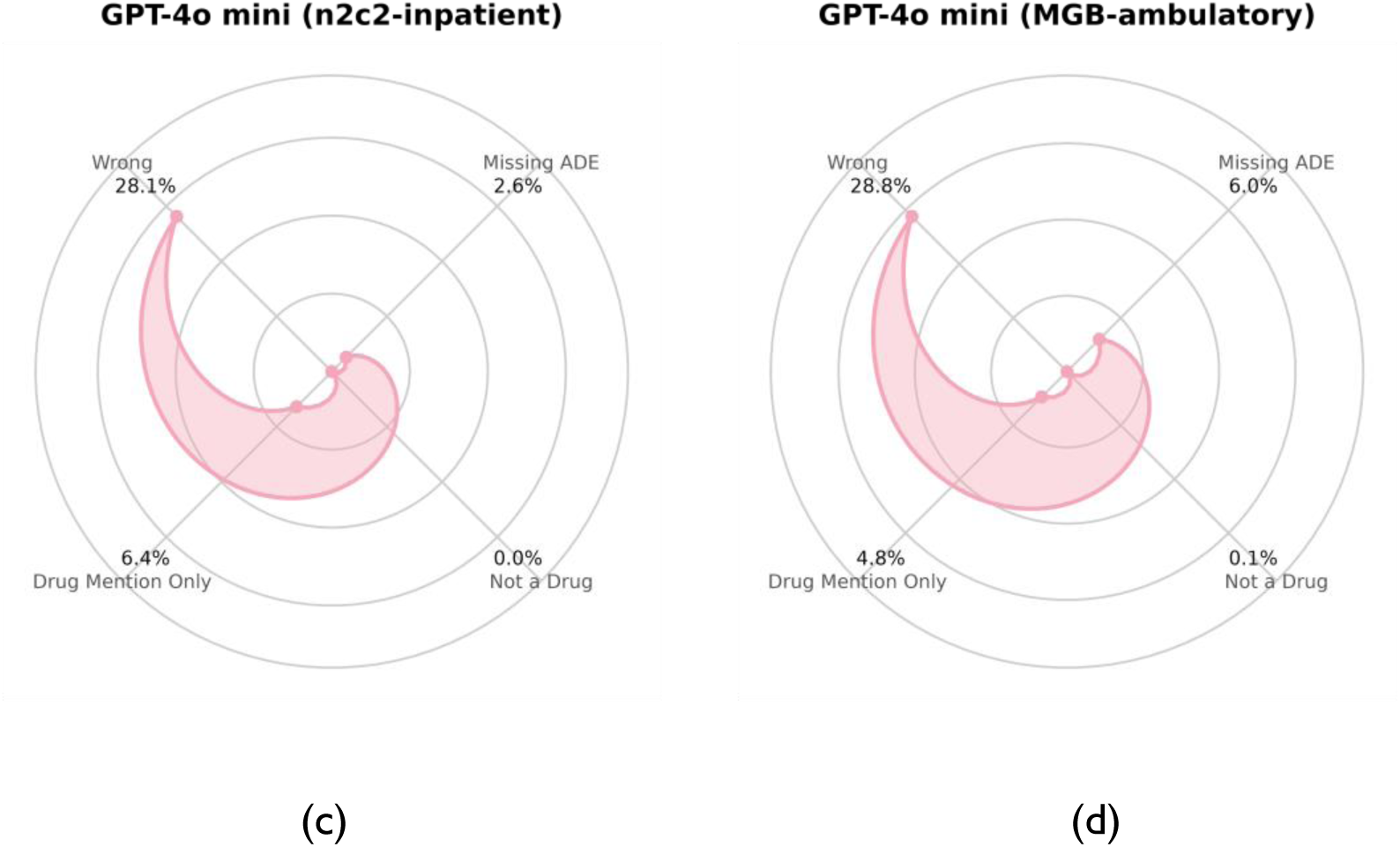
Error analysis of GPT-4o and GPT-4o-mini across two datasets. Radar plots display the distribution of error types, including wrong predictions, missing adverse events (ADEs), drug mention only, and “not a drug” errors. Results are shown for (a) GPT-4o on the n2c2-inpatient dataset, (b) GPT-4o on the MGB-ambulatory corpus, (c) GPT-4o-mini on the n2c2-inpatient dataset, and (d) GPT-4o-mini on the MGB-ambulatory corpus.

### Error Analysis

The models demonstrated strong performance in identifying ADEs when the clinical context was explicit and well-structured. For example, they correctly associated olanzapine with mild parkinsonian gait when the documentation suggested a medication-related etiology. The models also performed well in more complex scenarios, such as linking warfarin, metoprolol, and sacubitril/valsartan to a rash in the context of multiple newly initiated medications and a worsening dermatologic condition. Furthermore, they demonstrated the capacity to infer ADE-Drug associations from broader descriptions, exemplified by identifying captopril as a neurohormonal agent contributing to hypotension.

Despite these strengths, several limitations were observed. The models frequently misclassified pre-existing allergies as new ADEs, as illustrated by the incorrect attribution of severe reactions to doxycycline, ciprofloxacin, and cephalexin despite their documentation under known allergies. In addition, they often conflated therapeutic indications with adverse effects, such as labeling aspirin and clopidogrel as causing “blood clot prevention,” or erroneously identifying allergy therapies including diphenhydramine, prednisone, loratadine, and fexofenadine as causative agents for allergic reactions. Clostridium difficile cases are challenging because antibiotics may be both the cause of (non-exclusive in differential diagnosis) and the treatment for that indication. In some instances, the models recognized drug names but failed to establish any corresponding ADE, suggesting limitations in contextual interpretation.

Figure 3 presents an error analysis of GPT-4o and GPT-4o-mini across both the n2c2-inpatient and MGB-ambulatory corpus. For GPT-4o, most errors were wrong predictions, although the model produced relatively fewer missing ADEs and “not a drug” errors, particularly in the n2c2-inpatient dataset (Figure 3a). When applied to the MGB-ambulatory corpus, GPT-4o exhibited a higher number of wrong predictions (17.0%) and missing ADEs (Figure 3b). GPT-4o-mini showed a similar error profile but with a greater volume of wrong predictions (28.8%) overall, especially in the MGB-ambulatory corpus (Figure 3d). Across both models, “not a drug” errors were comparatively rare.

Figure 4 illustrates differences in the top frequency ADE-Drug pairs between the gold standard and model predictions. The gold standard (Figure 4a) highlights clinically well-recognized associations, including *lisinopril : angioedema*, *penicillin : rash*, and *heparin : thrombocytopenia*. GPT-4o (Figure 4b) reproduced several of these key signals, demonstrating partial alignment with the reference set, while also suggesting plausible but less established associations, such as *oxycodone : constipation*. By contrast, GPT-4o-mini (Figure 4c) captured some overlapping pairs but showed greater divergence, prioritizing ADE relations like *bactrim: rash* that were absent from the gold standard, as these are indications for treatment. Together, these comparisons show that GPT-4o aligns more closely with established evidence, whereas GPT-4o-mini produces a broader but less specific signal profile.

**Figure 4.**
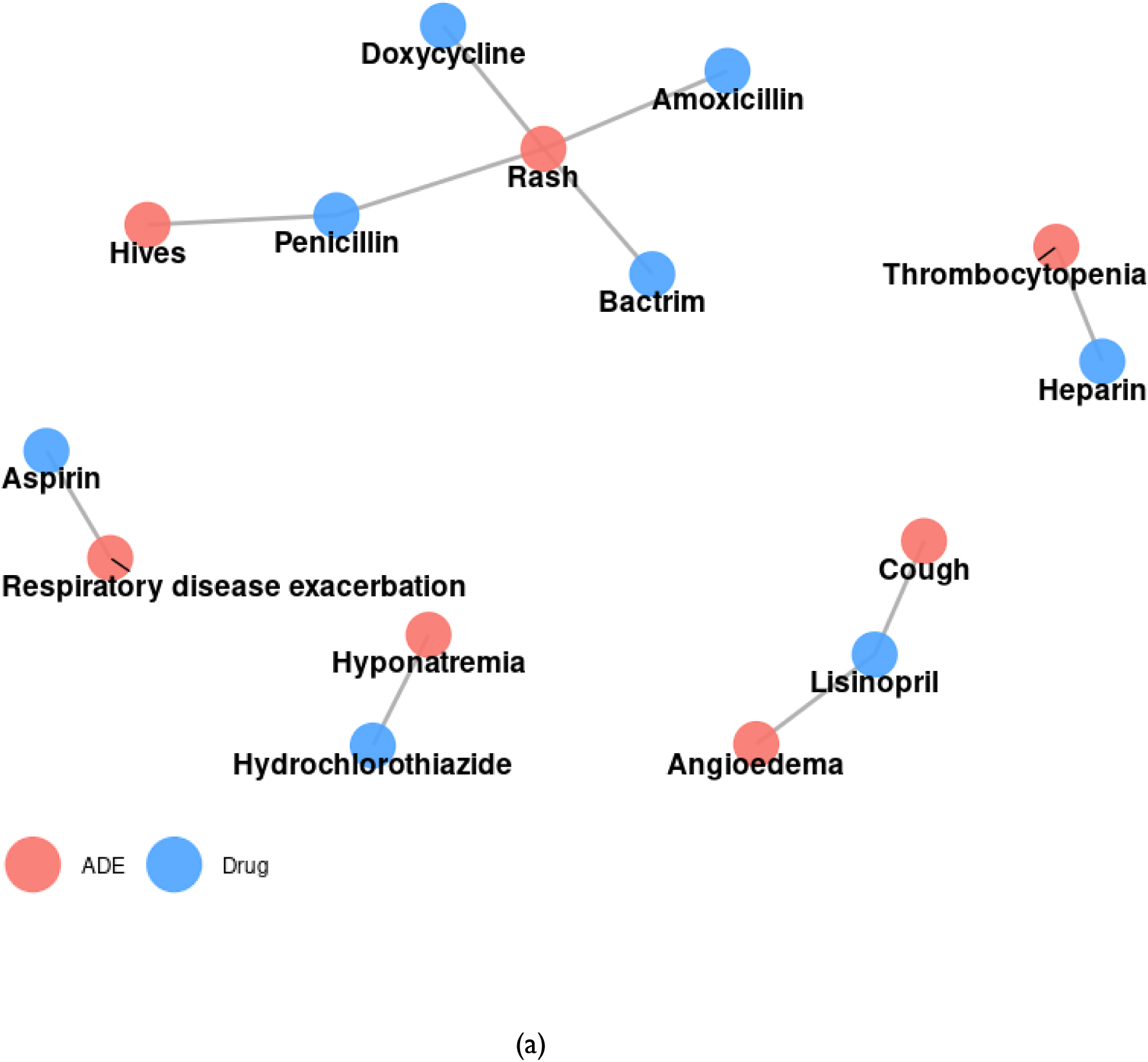
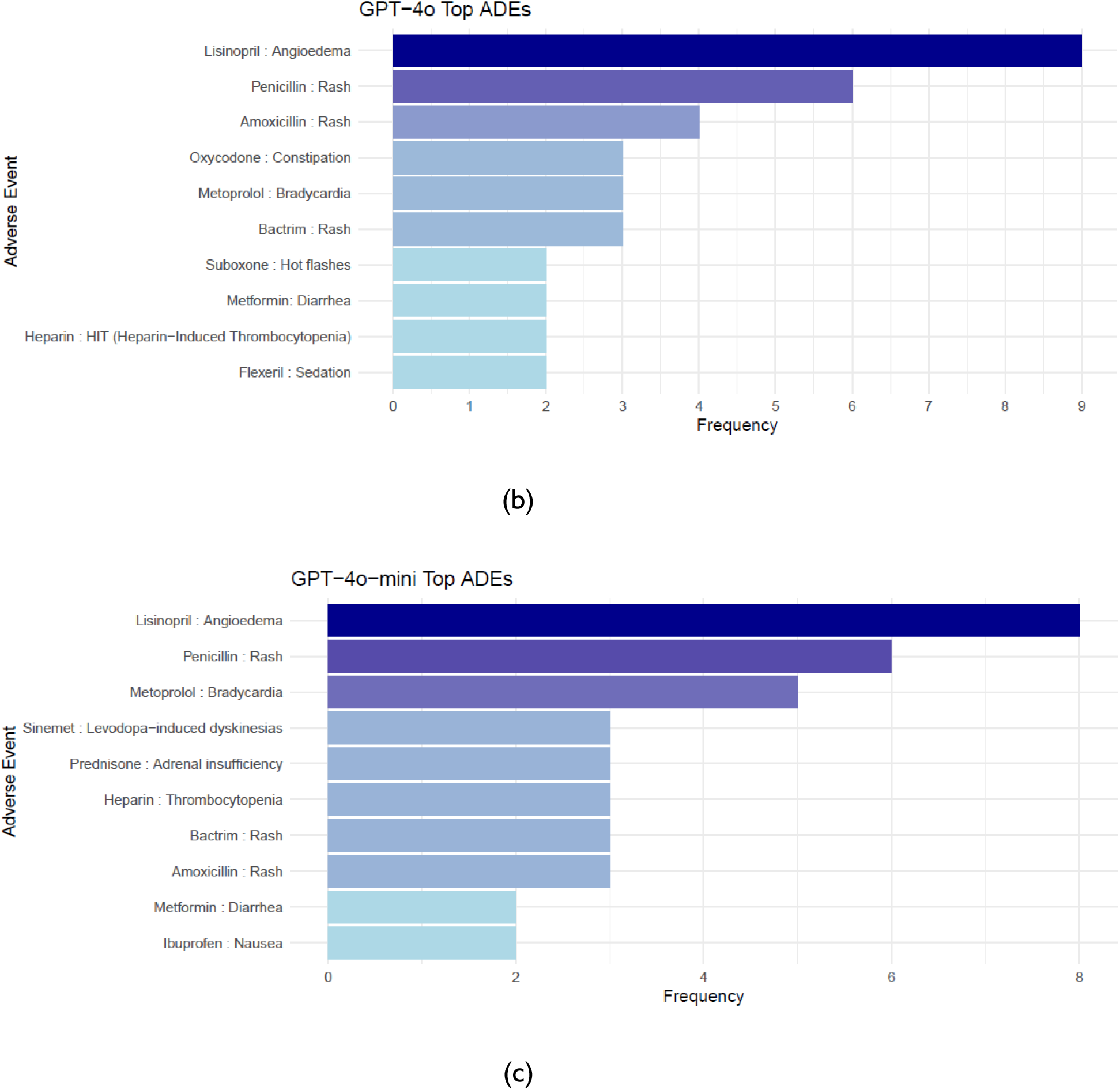
Comparison of top ADE-Drug pairs across gold standard and model predictions. (a) Network graph of the top ADE-Drug pairs in the gold standard reference set, highlighting well-recognized associations such as penicillin : rash, lisinopril : angioedema, and heparin : thrombocytopenia. (b) Barplot of the top ADE-Drug pairs predicted by GPT-4o, which reproduced several gold standard associations while also identifying additional signals, including oxycodone : constipation. (c) Barplot of the top ADE-Drug pairs predicted by GPT-4o-mini, which overlapped with some gold standard associations but also prioritized distinct signals, such as bactrim: rash.

## Discussion

### Principal Results

This study is one of the first to develop a comprehensive approach to identification of ADEs in outpatient notes. EHR clinical notes provided a robust data source for ADE detection. Our main finding is that while both models demonstrate limitations on complex, real-world text in terms of precision in ADE-Drug identification, GPT-4o provides more consistent performance across datasets, whereas GPT-4o-mini tends to generate higher error counts. ADE-Drug identification remains a challenge because the models are more than eager to return some sort of text to the question posed, even if it is wrong. Most of the wrong responses tended to be from the model returning an existing allergy (i.e., not an active ADE from the current visit), an indication (reason for use), or other hallucinations.

### Strengths

This study makes several novel contributions to the ADE extraction literature. We present a comprehensive evaluation of multiple state-of-the-art and recently released large language models, including fine-tuning strategies and low-rank adaptation (LoRA), which have not yet been widely studied for ADE extraction from clinical text. Most importantly, this is the first study to systematically examine ADE extraction in a large corpus of both private and public clinical notes, a setting that differs substantially from prior work focused on biomedical literature or social media (37–41). The scale of our annotated datasets meets established reporting quality guidelines for NLP studies, which recommend a minimum of 200 annotated documents, and the ambulatory corpus is large by the standards of manually annotated clinical NLP datasets (42). As ambulatory notes reflect real-world outpatient care, where most medications are prescribed and adverse events are first documented, our findings provide clinically meaningful evidence for the feasibility of deploying LLM-based ADE surveillance in routine clinical practice. In addition, by performing stratified analyses across urgent care and acute care settings, we demonstrate how model performance varies with clinical acuity and documentation context, strengthening the ecological validity and care-setting–specific interpretability of our results. Through focusing on ADE-Drug relationship classification, an underexplored but clinically critical task, and isolating relationship reasoning from upstream entity recognition variability, our evaluation provides clear insight into the real-world potential and limitations of LLMs for outpatient pharmacovigilance and clinical safety surveillance.

### Limitations

As the n2c2-inpatient dataset is before the training cutoff for the LLM models, it is possible that these models have been contaminated by including the n2c2-inpatient testing set within the LLM foundation model training data, especially as n2c2-inpatient utilized MIMIC III notes. We do not have access to the training data and thus cannot confirm in a white box manner. No black box testing was done on any of the models to try to re-create the n2c2-inpatient testing set using the LLMs, though using the n2c2-inpatient data in LLM foundation models would seem to violate terms of use.

Additionally, we limited the input text to a 225-character window surrounding annotated ADE-Drug spans to reduce noise, which may omit relevant information. In real-world deployment, gold-standard–derived snippets would be replaced by an upstream retrieval component (e.g., RAG), which may introduce modest performance degradation. Based on prior evidence that drug NER performance saturates at approximately 90–95% F1 due to label noise, we estimate a worst-case end-to-end F1 reduction of roughly 5–10% when using automated retrieval. However, as most ADE–Drug relations are expressed within one to two sentences of the relevant entities, clinically relevant context is unlikely to be systematically missed as other nearby entities would be identified, and thus missing drug concepts would be included in the snippets. While newer LLMs support longer context windows, increased context length does not necessarily improve accuracy and may introduce additional noise, consistent with prior findings that short-range context is often optimal for relation extraction. LLM performance may be sensitive to variation in clinical writing style, documentation practices, and note templates, including vendor-specific or structured EHR elements (e.g., Epic SmartPhrases), which can influence how ADEs are expressed in text. As our evaluation was conducted using data from a single healthcare network, the applicability of our findings to other EHR vendors or differently structured clinical notes remains an open question. This research was conducted at a single tertiary referral academic center which may limit the generalizability of the findings. However, our center has both large general and extensive subspecialty ambulatory practices, which has ensured that our study sample includes a wide variety of ADE cases, including rare, severe ones.

### Comparison with Prior Work

While overall system performance of the top-performing system in the n2c2-inpatient challenge was reported to be 94.18% F1 for concept extraction, 96.3% F1 for relation classification, and 89.05% F1 for end-to-end performance, these metrics are largely related to excellent performance on the extraction of concepts related to drug signature information(23). Performance on the ADE-Drug label was reported to be 47.55% label-wise F1 on the n2c2-inpatient dataset for the top-performing system(23). While not directly comparable due to a difference between span-level and document-level metrics, GPT-4o achieved 53.0% F1 on the n2c2-inpatient dataset, whereas GPT-4o-mini reached only 33.0%. These traditional NLP systems often have issues that LLMs may not with respect to undetected negations (e.g., “did take a dose of [drugs] prior to the procedure but the rash had proceeded her taking the medications”), compound sentence patterns (e.g., “[symptoms] s/p receiving [drug]”), context switches (e.g., going from discussing an ADE to discussing an unrelated drug “using [drug] with some benefit” then back to discussing the ADE), scoping attributable to the loss of original note structure (e.g., spacing, punctuation, paragraphs, sections), misspellings, abbreviations, and inclusion of broader/vague substances in the ground truth (e.g., drug classes, supplements, “drug”) that are not in the standard medication dictionary.

Our study also identified critical areas for improvement in ADE detection. We found that indications may have been misannotated by NLP and attributed as an ADE; in the real-world context, this could be frustrating to a tool’s clinician users. One approach to mitigate this issue would be to add annotations of indication to the annotated dataset, from which NLP could utilize joint modeling. Prior research has shown a 3% F1-measure improvement when joint modeling of relations between medications and ‘signs or symptoms’ were used to jointly determine ADE and indication entities.^30^ Indication-Drug relationships were a common issue identified in both models responses in this study.

In our initial experimentation, we found that the LLMs did not do well with noisy input, and that pruning input to a relevant short-range context improved performance. This finding provides additional evidence to support Dandala et al.’s observation that long-range context created noise while short-range context tended to be more accurate (43). The optimization of allowing long-vs-medium-vs-short-range relationships is complex, and how best to teach models to disregard irrelevant information remains an open research question. In practice, in an end-to-end pipeline, a Retrieval-Augmented Generation (RAG) component or LLM to identify relevant context would replace our gold standard pre-processing step. We used this pre-processing step for convenience from having an annotated gold standard so that performance of the tools would be comparable (i.e., based on the same snippets of text), but real-world applications are unlikely to be fully annotated.

## Conclusions

This study shows that generative large language models enable reliable identification of ADE-Drug relationships from unstructured clinical notes. Through scalable and accurate extraction of safety signals, these models represent an important step forward for pharmacovigilance, supporting earlier detection of adverse events and strengthening patient safety efforts.

## Data Availability

The n2c2 data may be available from i2b2 via a Data Use Agreement. The Mass General Brigham data generated and analyzed during the current study are not publicly available due to organizational patient privacy policies. Any request to access the Data will need to be reviewed and approved by Mass General Brigham. Researchers will need to provide evidence of IRB approval for their study. For the approved studies, data will be released via a Data Use Agreement.

## Acknowledgments

We thank Abigail Salem, Lynn Volk, Suzanne V. Blackley, M.S., Amol Rajmane, M.D., M.S., M.B.A., Jane Snowdon, and Winnie Felix for administrative assistance. We thank Amy Boughan, N.P., Cecilia Sarkissian, M.D., Jean Yeung, M.D., M.SC., Kathleen Sheehan, R.N., Kathleen Tobin, R.N., Kit Chan, M.D., Margaret Orrick, R.N., Pracha Peter Eamranond, M.D., M.P.H., Sadia Hussain, M.D., Theresa Love, R.N., Yih-Chieh Chen, M.D., and Heba Edrees, Pharm.D. for help annotating the clinical notes. We thank Stuart Lipsitz, Sc.D., Phuc Tran, Paul L Felt, PhD, Brendan Bull, Brett R. South, Ph.D., Mario J. Lorenzo, Ph.D., Deb Angst, and Henry J. Feldman, M.D. for assistance with the larger study from which this effort evolved.

## Author Contributions

All authors contributed substantially to the conception and design of this work, its data analysis and interpretation, and helped draft and revise the manuscript. All authors are accountable for the integrity of this work.

CRedit: **Joseph M. Plasek:** Conceptualization, Data curation, Formal analysis, Investigation, Methodology, Project Administration, Software, Validation, Visualization, Writing – original draft, Writing – review & editing. **Yiming Li:** Conceptualization, Formal analysis, Methodology, Software, Validation, Visualization, Writing – original draft, Writing – review & editing. **Mary G. Amato:** Data curation, Validation, Writing – review & editing. **Dinah Foer:** Data curation, Writing – review & editing. **Diane L. Seger:** Data curation, Writing – review & editing, **Shayma Alzaidi:** Data curation, Writing – review & editing. **Huiyuan Zhou:** Visualization, Writing – review & editing. **Gretchen Purcell Jackson:** Funding acquisition, Writing – review & editing. **David W. Bates:** Funding acquisition, Writing – review & editing. **Li Zhou:** Conceptualization, Funding acquisition, Methodology, Project administration, Resources, Supervision, Writing – review & editing.

## Ethics Approval and Consent to Participate

This study complied with the World Medical Association Declaration of Helsinki on Ethical Principles for Medical Research Involving Human Subjects. It was reviewed by and approved by the Mass General Brigham Institutional Review Board (IRB) (reference number: 2019P002998). The IRB waived the requirement of consent for this study.

